# Self-Care Competence and AI-Supported Learning as Predictors of Enhanced Clinical Decision-Making Skills Among Nurses

**DOI:** 10.64898/2026.05.02.26352291

**Authors:** Caleb Onah, Adawa Ibrahim Haruna

## Abstract

Clinical decision-making is a critical competency for nurses, particularly in resource-constrained healthcare systems where frontline practitioners must integrate clinical knowledge, judgment, and contextual constraints to ensure optimal patient outcomes. Although prior research highlights the benefits of artificial intelligence (AI)-supported learning and individual competencies, it largely assumes a direct relationship between technological support and decision quality, overlooking the cognitive-regulatory mechanisms through which such effects occur. This study addresses this gap by examining self-care competence as a mediating pathway linking perceived AI-based learning support to enhance clinical decision-making among nurses in Benue State, Nigeria. A descriptive cross-sectional design was employed, with data collected from 600 registered nurses across public and private healthcare facilities using the Self-Care Competence Scale (SCCS), AI-Based Learning Support Scale (PAILS) and the Clinical Decision-Making Scale (CDMNS). Data were analyzed using structural equation modelling (SEM) to test direct and indirect relationships, complemented by bootstrapped mediation analysis and rigorous assessment of common method bias through Harman’s single-factor test and full collinearity variance inflation factors, ensuring robustness of the findings. Results indicated moderately high levels of self-care competence, perceived AI-based learning support, and enhance clinical decision-making skills. Self-care competence and AI-based learning support significantly predicted clinical decision-making, with self-care competence partially mediating this relationship and the model explaining 58 percent of the variance. The findings extend theory by demonstrating that AI-supported learning enhances enhance clinical decision-making not directly, but through nurses’ cognitive and psychological readiness, positioning self-care competence as a central mechanism in evidence-based practice.

## Background

The increasing complexity of healthcare delivery systems globally has placed significant demands on nurses to demonstrate high levels of clinical decision-making skills. Enhance clinical decision-making is a fundamental component of nursing practice, involving the ability to assess patient conditions, interpret data, and implement appropriate interventions to ensure optimal patient outcomes. In contemporary healthcare settings, nurses are expected not only to provide routine care but also to engage in critical thinking and evidence-based practice in making clinical judgments (Zainal et al., 2026; Chinonyelum et al., 2025). This expectation is particularly important in developing countries such as Nigeria, where nurses constitute a large proportion of the healthcare workforce and often serve as the primary point of care in both urban and rural health facilities (Olabode et al., 2024; Onah et al., 2023).

In Nigeria, the healthcare system continues to face numerous challenges, including inadequate staffing, limited resources, and uneven distribution of healthcare professionals. These challenges increase the burden on nurses and require them to make timely and accurate clinical decisions under pressure. Studies have shown that nurses contribute up to 90% of primary healthcare services in low-resource settings, emphasizing the importance of their decision-making competence in improving patient outcomes (Olabode et al., 2024). However, despite their central role, there remains a gap between knowledge acquisition and the practical application of enhance clinical decision-making skills among nurses in Nigeria (Olabode et al., 2024).

Clinical decision-making is closely linked to clinical competence, which encompasses knowledge, skills, attitudes, and the ability to apply these effectively in patient care. Evidence from Nigerian studies indicates that clinical competence among nurses and nursing students is influenced by several factors, including training quality, exposure to clinical experiences, and self-assessment abilities (Chinonyelum et al., 2025). Furthermore, the use of standardized nursing care plans, which are intended to guide clinical decisions, has not been fully optimized in many Nigerian healthcare institutions due to implementation challenges and institutional limitations (Ojo et al., 2023). This suggests that nurses may lack structured support systems to enhance their decision-making processes.

Another critical factor that may influence enhance clinical decision-making among nurses is self-care competence. Self-care competence in this study, refers to the ability of nurses to maintain their physical, emotional, and psychological well-being. In the nursing profession, self-care is essential because nurses are frequently exposed to stressful working conditions, long shifts, and emotional demands associated with patient care. Research has shown that poor self-care practices among nurses can lead to burnout (Edmealem et al., 2024), reduced productivity, and impaired cognitive functioning, all of which negatively affect enhance clinical decision-making abilities. In the Nigerian context, where healthcare systems are often overstretched, the importance of self-care competence becomes even more pronounced. Nurses who are unable to manage stress effectively may struggle to make sound clinical judgments, thereby compromising patient safety.

In addition to self-care competence, technological advancements have introduced new dimensions to nursing practice, particularly through the integration of artificial intelligence (AI) in healthcare. AI-based learning support systems are increasingly being used to enhance knowledge acquisition, clinical reasoning, and decision-making among healthcare professionals. These systems provide real-time access to clinical guidelines, diagnostic tools, and decision-support algorithms, thereby assisting nurses in making more informed decisions (Ominyi et al., 2025). Although the adoption of AI in Nigeria is still emerging, there is growing recognition of its potential to improve healthcare delivery and professional development among nurses. Studies have highlighted that technology-driven learning approaches can improve nurses’ knowledge, confidence, and clinical skills, which are essential components of effective decision-making (Guerrero et al., 2024).

Despite the potential benefits of AI-based learning support, its utilization in Nigerian healthcare settings remains limited due to factors such as inadequate infrastructure, lack of training (Guerrero et al., 2024; Glauberman et al., 2023), and resistance to change. Moreover, there is limited empirical research examining how perceived AI-based learning support influences enhance clinical decision-making among nurses in specific regions such as Benue State. This gap in the literature underscores the need for studies that explore the role of emerging technologies in enhancing nursing practice within the Nigerian context.

Furthermore, participation in clinical decision-making processes among nurses in Nigeria has been found to vary across healthcare settings. While nurses are expected to collaborate with other healthcare professionals in decision-making, studies suggest that their level of involvement is sometimes restricted due to hierarchical structures and organizational policies (Dokuba & Eleke, 2021). This limited participation may hinder the development of critical thinking and decision-making skills, as nurses may not have sufficient opportunities to engage actively in clinical judgments (Zainal et al., 2026).

Given these challenges, there is a growing need to identify key predictors of enhance clinical decision-making skills among nurses. Self-care competence and perceived AI-based learning support represent two important variables that may significantly influence nurses’ ability to make effective clinical decisions. While self-care competence ensures that nurses are mentally and physically capable of performing their duties, AI-based learning support provides the necessary tools and resources to enhance their knowledge and clinical reasoning.

In Benue State, where healthcare facilities face similar challenges as other parts of Nigeria (Josiah et al., 2019), understanding these predictors is crucial for improving nursing practice and patient outcomes. The state’s healthcare system relies heavily on nurses to deliver essential services, particularly in rural areas where access to physicians may be limited. Therefore, enhancing enhance clinical decision-making skills among nurses in this region is vital for strengthening the overall healthcare system.

Thus, enhance clinical decision-making remains a critical skill for nurses, particularly in resource-constrained settings such as Nigeria. While existing literature has explored factors influencing clinical competence and decision-making, there is limited research focusing on the combined influence of self-care competence and perceived AI-based learning support. This study, therefore, seeks to fill this gap by examining how these variables predict enhance clinical decision-making skills among nurses in Benue State. By doing so, it aims to contribute to the growing body of knowledge on nursing practice and provide evidence-based recommendations for improving healthcare delivery in Nigeria.

### Objectives of the Study

The specific objectives are to:

1. Examine the influence of self-care competence on enhance clinical decision-making skills among nurses in Benue State.
2. Determine the influence of perceived AI-based learning support on enhance clinical decision-making skills among nurses in Benue State.
3. Assess the joint (combined) predictive influence of self-care competence and perceived AI-based learning support on enhance clinical decision-making skills among nurses in Benue State.

### Hypotheses

1. Self-care competence will significantly influence on enhance clinical decision-making skills among nurses in Benue State.
2. Perceived AI-based learning support will significantly influence enhance clinical decision-making skills among nurses in Benue State.
3. Self-care competence and perceived AI-based learning support will significantly and joint influence on enhance clinical decision-making skills among nurses in Benue State.

### Theoretical Framework: An Integrated EBP–Cognitive Load–TAM Perspective

This study is anchored in an integrated theoretical framework that combines Evidence-Based Practice (EBP), Cognitive Load Theory (CLT), and the Technology Acceptance Model (TAM) to provide a comprehensive explanation of enhance clinical decision-making among nurses. Rather than relying on a single theoretical lens, this integrative approach recognizes that enhance clinical decision-making is shaped by the interaction between external knowledge systems, cognitive capacity, and technology perception. Evidence-Based Practice (EBP) serves as the foundational framework, emphasizing that clinical decisions should be guided by the integration of the best available evidence, professional expertise, and patient needs (Naz & Ganaie, 2023). Within this study, EBP represents the external knowledge domain, which is increasingly supported by AI-driven learning tools that provide access to real-time clinical information.

**Diagram 1:**
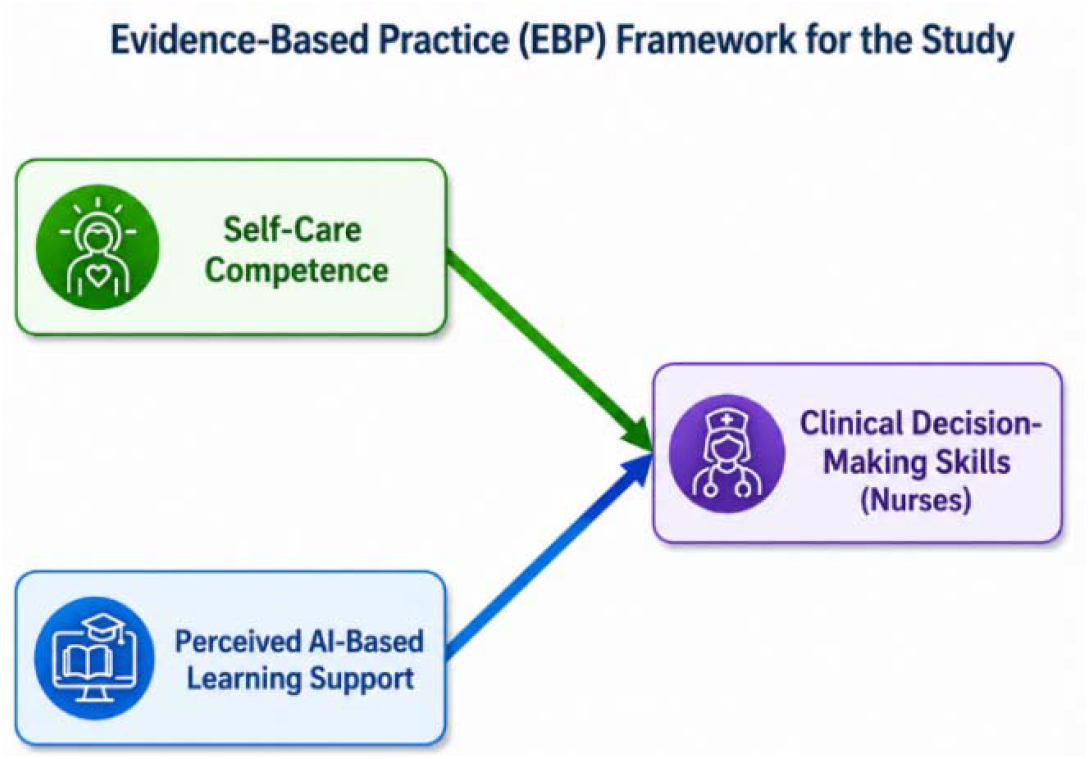
Evidence-Based Practice (EBP)

EBP theory posits that effective enhance clinical decision-making is a systematic process involving the acquisition, evaluation, and application of evidence rather than reliance on intuition alone. The adoption of EBP among nurses has been shown to improve the quality and accuracy of clinical decisions, particularly in resource-constrained settings (Olabode et al., 2024). This is especially relevant in Benue State, where nurses often function as frontline healthcare providers and must depend on both internal competencies and external knowledge systems to deliver effective care. However, while EBP explains what constitutes effective decision-making, it does not fully account for variations in nurses’ capacity to process and apply evidence under conditions of stress or cognitive demand.

To address this limitation, Cognitive Load Theory (CLT) is incorporated to explain the cognitive mechanisms underlying clinical decision-making. CLT posits that human cognitive capacity is limited, and excessive mental load—arising from stress, fatigue, or complex task demands—can impair information processing and decision quality. In nursing contexts, enhance clinical decision-making requires sustained attention, working memory, and emotional regulation (East African Health Research Commission, 2024). When cognitive load exceeds manageable levels, decision accuracy declines. Within this framework, self-care competence functions as a cognitive-regulatory resource that helps reduce mental strain, enhance attentional control, and sustain cognitive performance. Thus, self-care competence enables nurses to more effectively engage in evidence-based reasoning by preserving the cognitive capacity required to interpret and apply clinical information.

In parallel, the Technology Acceptance Model (TAM) is integrated to explain how nurses engage with AI-based learning systems as sources of external evidence. TAM posits that individuals’ adoption of technology is primarily determined by perceived usefulness and perceived ease of use. In this study, perceived AI-based learning support reflects these dimensions, influencing whether nurses utilize AI-driven tools in their clinical practice. AI-based systems provide real-time access to guidelines, diagnostic support, and evidence-based resources, thereby operationalizing the principles of EBP in modern healthcare environments (Edmealem et al., 2024). However, the mere availability of such tools does not guarantee their effective use. Rather, their impact depends on nurses’ perceptions and willingness to integrate them into decision-making processes, as explained by TAM.

The integration of these three theories provides a mechanistic understanding of clinical decision-making. EBP explains the role of external evidence systems (AI tools), TAM explains the behavioral and perceptual factors influencing engagement with these systems, and CLT explains the cognitive capacity required to process and apply the information provided. Together, they suggest that AI-based learning support enhances enhance clinical decision-making not directly, but through its interaction with nurses’ cognitive readiness and psychological well-being. In this sense, self-care competence becomes a critical pathway through which technological inputs are translated into effective clinical judgments.

In the Nigerian healthcare context, the application of this integrated framework is particularly important. The implementation of EBP remains constrained by limited resources, inadequate training, and organizational barriers (Ojo et al., 2023). As a result, nurses often rely on alternative mechanisms, such as AI-based learning tools, to access up-to-date clinical information. However, variability in digital literacy and infrastructure affects the extent to which these tools are utilized. While TAM explains differences in technology adoption and CLT explains variations in cognitive functioning, EBP provides the overarching structure that integrates both human and technological dimensions into a unified model of clinical decision-making.

Finally, this integrated framework emphasizes that effective enhance clinical decision-making emerges from the alignment between internal capacity and external support systems. Nurses must possess sufficient cognitive and emotional resources (as explained by CLT and reflected in self-care competence) while also having access to and willingness to use evidence-based technological tools (as explained by TAM and operationalized through AI-based learning support). When these elements are aligned, nurses are better positioned to engage in evidence-based practice and deliver high-quality care (Olabode et al., 2024). Additionally, enhance clinical decision-making is a dynamic process shaped by continuous learning, adaptation, and critical thinking (Zhang et al., 2025). In resource-limited settings such as Benue State, this integrated EBP–CLT–TAM framework provides a robust and contextually relevant explanation for enhancing enhance clinical decision-making among nurses.

## Methods

### Design and Setting

A descriptive cross-sectional research design was used to conduct the current study. This design enabled the assessment of relationships among self-care competence, perceived AI-based learning support, and enhance clinical decision-making skills among nurses at a single point in time without manipulation of variables. The study followed the Strengthening the Reporting of Observational Studies in Epidemiology (STROBE) guidelines for reporting observational research studies.

The study was conducted in selected public and private healthcare facilities in Benue State, Nigeria. Benue State is located in the North-Central region of Nigeria and comprises a mix of tertiary, secondary, and primary healthcare institutions serving both urban and rural populations. These facilities provide a wide range of healthcare services and depend heavily on nurses for patient care delivery and clinical decision-making. The selected institutions included general hospitals, teaching hospitals, and primary healthcare centres, all of which manage diverse patient populations under varying resource conditions.

### Participants and Sampling

The multistage sampling approach with a pragmatic recruitment strategy was employed to enhance representativeness while accommodating field constraints. In the first stage, healthcare facilities were purposively selected to reflect the diversity of care levels in Benue State, including primary healthcare centres, general hospitals, and tertiary institutions. This ensured adequate coverage of varying clinical environments and decision-making demands. In the second stage, proportionate sampling was applied within each selected facility to recruit registered nurses across departments and experience levels. Within these strata, participants were recruited using an availability-based approach due to workforce schedules and institutional access limitations.

Importantly, the inclusion of nurses across multiple facility types, experience levels, and educational backgrounds enhances the heterogeneity and analytical generalizability of the sample. However, it is acknowledged that the use of an availability-based recruitment component may introduce selection bias, potentially limiting external validity. To mitigate this, efforts were made to include a large and diverse sample (N = 600) across different healthcare levels.

To determine the required sample size for structural equation modelling (SEM), a priori power analysis was conducted. Considering an anticipated medium effect size (0.15), an alpha level of 0.05, statistical power of 0.80, and the complexity of the proposed model with latent constructs, a minimum sample size of 200 participants was recommended to ensure adequate model stability and reliable parameter estimation. This recommendation aligns with established SEM guidelines, which suggest a minimum sample size ranging from 200 to 400 depending on model complexity.

The demographic characteristics of the participants provide a clear picture of the nursing workforce represented in this study.

The age distribution shows that the majority of respondents were within the active working age group. Most nurses (40.0%) were between 30–39 years, followed by those aged 20–29 years (30.0%). Nurses aged 40–49 years constituted 20.0%, while only 10.0% were 50 years and above. This indicates that the sample was largely composed of young to middle-aged professionals, suggesting a workforce that is still actively engaged in clinical practice and likely adaptable to new learning approaches such as AI-based support systems.

In terms of gender, the sample was predominantly female (65.0%), with males accounting for 35.0%. This reflects the general gender distribution in the nursing profession, where females typically constitute the majority, especially in developing countries like Nigeria.

Regarding marital status, most participants were married (60.0%), while 35.0% were single and a smaller proportion (5.0%) were divorced or widowed. This distribution suggests that a significant number of respondents may be balancing professional responsibilities with family obligations, which could influence self-care practices and decision-making processes.

Educational qualification data revealed that half of the participants (50.0%) held a Bachelor of Nursing Science (BNSc) degree, while 30.0% possessed diploma qualifications (RN/RM), and 20.0% had postgraduate degrees. This indicates a relatively well-educated sample, with a substantial proportion having advanced training, which is important for clinical reasoning and decision-making competence.

The distribution of years of professional experience shows a balanced mix of early-career and experienced nurses. The largest group (35.0%) had 5–10 years of experience, followed by those with less than 5 years (25.0%) and 11–20 years (25.0%). Nurses with over 20 years of experience constituted 15.0%. This diversity in experience levels enhances the representativeness of the sample and provides a broader perspective on enhance clinical decision-making skills.

Finally, with respect to the type of healthcare facility, the majority of participants were drawn from general hospitals (45.0%), followed by teaching/tertiary hospitals (35.0%), and primary healthcare centres (20.0%). This suggests that most respondents operate in secondary and tertiary care settings, where enhance clinical decision-making demands are often higher due to the complexity of cases managed.

Overall, the demographic profile indicates a predominantly female, moderately experienced, and well-educated nursing workforce, largely concentrated in hospital-based settings. This composition is appropriate for examining self-care competence, perceived AI-based learning support, and enhance clinical decision-making skills, as these factors are particularly relevant in such professional contexts. Although convenience sampling was used due to access constraints, the relatively large sample size (N = 600) and inclusion of nurses across different healthcare settings (primary, secondary, and tertiary institutions), educational levels, and years of experience enhance the diversity and improve the representativeness of the sample. This diversity increases the robustness of the findings and supports their applicability within similar healthcare contexts.

### Data collection

Data were collected over a three-month period (between January – March, 2026). Following ethical approval from the appropriate institutional review board – Benue State Ministry of Health and Human Service, Benue State Hospital Management Borad, permission was obtained from the management of selected healthcare facilities along with a detailed explanation of the study objectives and significance. The questionnaire was distributed both in paper format and electronically via online platforms such as WhatsApp to enhance participation. The survey link was shared with eligible nurses along with an invitation explaining the purpose of the study and assuring voluntary participation and confidentiality. Measures were taken to prevent duplicate responses in the online survey. Participants completed the questionnaire anonymously.

Administrative permission was also obtained from participating healthcare facilities and centres. Participants were informed that there were no anticipated risks associated with participation and that they could withdraw at any time without penalty. Privacy and confidentiality were strictly maintained, with no identifying information collected. All responses were kept anonymous and used solely for research purposes. Written and recorded verbal informed consent was obtained from all participants before completion of the questionnaire. Data was collected using the following tools:

1. **Sociodemographic data:** This section included age, sex, marital status, level of education, years of professional experience, and type of healthcare facility.
2. **Self-Care Competence Scale (SCCS):** The SCCS was originally developed by Schmitt (2013) and later adapted for healthcare professionals in subsequent nursing research contexts. The scale comprises multiple Likert-type items rated on a 5-point scale (1 = strongly disagree to 5 = strongly agree), with higher scores indicating greater self-care competence across physical, emotional, and psychological domains. The SCCS has demonstrated strong internal consistency, with Cronbach’s alpha coefficients ranging from 0.78 to 0.92 across different populations. In advanced psychometric evaluations, composite reliability (CR) values have been reported above 0.80, confirming adequate internal consistency at the construct level. Construct validity has been well established through both exploratory and confirmatory factor analyses. Convergent validity is supported by Average Variance Extracted (AVE) values exceeding the recommended threshold of 0.50, indicating that the items adequately represent the latent construct. Discriminant validity has also been confirmed using the Heterotrait–Monotrait (HTMT) ratio, with values below 0.85, demonstrating that SCCS is empirically distinct from related constructs such as burnout and job stress. CFA results from prior validation studies indicate good model fit indices, typically within acceptable thresholds: RMSEA ≈ 0.05–0.07, CFI ≈ 0.90–0.95, TLI ≈ 0.90–0.94, GFI ≈ 0.88–0.92, and AGFI ≈ 0.85–0.90. These indices confirm the factorial stability of the scale across healthcare samples. Item-total correlations generally range from 0.40 to 0.75, indicating adequate item discrimination. Factor loadings typically fall between 0.60 and 0.85, suggesting strong relationships between observed variables and the latent construct. Overall, the SCCS demonstrates robust psychometric properties, including high reliability, strong construct validity, and acceptable factorial structure, making it suitable for assessing self-care competence among nurses in both clinical and research settings.
3. **AI-Based Learning Support Scale (PAILS):** The scale is adapted from the foundational work of Davis (1989) on TAM, with subsequent modifications in AI and e-learning research contexts (2018–2024). The instrument includes multiple Likert-scale items (1 = strongly disagree to 5 = strongly agree) assessing four domains: perceived usefulness, perceived ease of use, accessibility, and effectiveness (Zhang et al., 2021a). Higher scores reflect stronger perceived AI-based learning support. The PAILS demonstrates excellent internal consistency, with Cronbach’s alpha values ranging from 0.85 to 0.94. Composite reliability indices are typically above 0.85, confirming strong reliability at the latent construct level. Convergent validity is supported by AVE values exceeding 0.50, while discriminant validity is confirmed through HTMT ratios below 0.85. The scale also shows strong nomological validity, as it correlates significantly with technology usage, learning outcomes, and decision-making performance. CFA results indicate excellent model fit: RMSEA ≈ 0.04–0.06, CFI ≈ 0.93–0.97, TLI ≈ 0.92–0.96, NFI ≈ 0.90– 0.95, and SRMR ≈ 0.03–0.06. These indices demonstrate that the measurement model aligns well with empirical data. Factor loadings typically range from 0.65 to 0.90, indicating strong item representation of the latent construct. Item discrimination indices fall between 0.45 and 0.80, confirming that items effectively differentiate between respondents with varying levels of perceived AI support. The PAILS exhibits excellent psychometric robustness, including high reliability, strong construct validity, and superior model fit indices, making it a reliable instrument for assessing perceived AI-based learning support in healthcare contexts (Zhang et al., 2021b).
4. **Clinical decision-making in Nursing Scale (CDMNS):** The scale was developed by Judith M. Jenkins (1985). The CDMNS consists of 40 items across four domains: search for alternatives, canvassing of objectives and values, evaluation and re-evaluation of consequences, and search for information. Items are rated on a 5-point Likert scale, with higher scores indicating stronger enhance clinical decision-making ability. The CDMNS demonstrates excellent reliability, with Cronbach’s alpha coefficients ranging from 0.83 to 0.97. Subscale reliabilities typically exceed 0.80, while composite reliability values are above 0.85, confirming high internal consistency. Construct validity has been extensively established across diverse nursing populations. Convergent validity is supported by AVE values above 0.50, while discriminant validity is confirmed through HTMT ratios below 0.85. Criterion-related validity has also been demonstrated through significant correlations with clinical competence and critical thinking measures. CFA findings indicate strong model fit: RMSEA ≈ 0.05–0.08, CFI ≈ 0.90–0.95, TLI ≈ 0.90–0.94, GFI ≈ 0.88–0.92, and NFI ≈ 0.89–0.93. These values support the four-factor structure of the scale. Factor loadings range from 0.60 to 0.88, indicating strong item contributions. Item-total correlations typically fall between 0.45 and 0.78, reflecting good discrimination power. The scale has demonstrated stability across different cultural and clinical contexts. Thus, the CDMNS remains one of the most psychometrically sound instruments for assessing enhance clinical decision-making in nursing, with strong reliability, well-established validity, and consistent factorial structure across studies.

### Data analysis plan

All analyses were conducted using R software version 4.2.2. Descriptive statistics were used to summarize participant characteristics. Continuous variables such as age and years of experience were presented using means and standard deviations as shown in table 1, while categorical variables such as sex, marital status, and education level were summarised using frequencies and percentages.

**Table 1:**
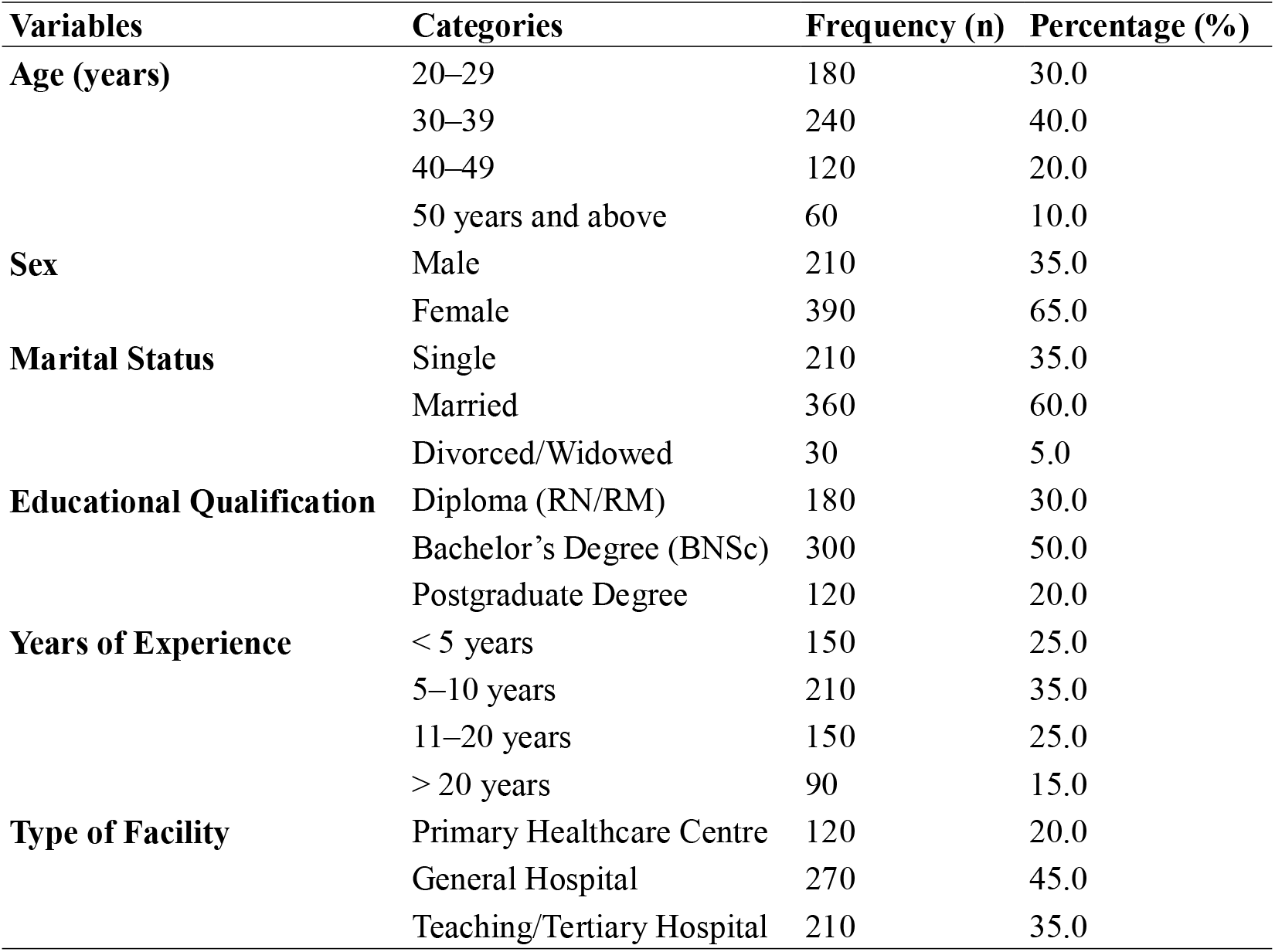
Sociodemographic Characteristics of Participants (N = 600)

| Variables | Categories | Frequency (n) | Percentage (%) |
| --- | --- | --- | --- |
| **Age (years)** | 20–29 | 180 | 30.0 |
|  | 30–39 | 240 | 40.0 |
|  | 40–49 | 120 | 20.0 |
|  | 50 years and above | 60 | 10.0 |
| **Sex** | Male | 210 | 35.0 |
|  | Female | 390 | 65.0 |
| **Marital Status** | Single | 210 | 35.0 |
|  | Married | 360 | 60.0 |
|  | Divorced/Widowed | 30 | 5.0 |
| **Educational Qualification** | Diploma (RN/RM) | 180 | 30.0 |
|  | Bachelor’s Degree (BNSc) | 300 | 50.0 |
|  | Postgraduate Degree | 120 | 20.0 |
| **Years of Experience** | < 5 years | 150 | 25.0 |
|  | 5–10 years | 210 | 35.0 |
|  | 11–20 years | 150 | 25.0 |
|  | > 20 years | 90 | 15.0 |
| **Type of Facility** | Primary Healthcare Centre | 120 | 20.0 |
|  | General Hospital | 270 | 45.0 |
|  | Teaching/Tertiary Hospital | 210 | 35.0 |

Structural equation modelling (SEM) was employed to examine the relationships among self-care competence, perceived AI-based learning support, and enhance clinical decision-making skills. The analysis was conducted using the “lavaan” package in R. The measurement model was first assessed to confirm the validity of the latent constructs, followed by the structural model to test the hypothesized relationships among variables. Model fit was evaluated using multiple indices, including Chi-square statistic/degree of freedom (χ^2^/df), Comparative Fit Index (CFI), Tucker-Lewis Index (TLI), Root Mean Square Error of Approximation (RMSEA), and Standardized Root Mean Square Residual (SRMR). A χ^2^/df value less than 5 indicated acceptable fit, while CFI and TLI values greater than 0.90 indicated good fit. RMSEA values ≤ 0.05 indicated good fit, while values ≥ 0.10 indicated poor fit. SRMR values below 0.08 were considered acceptable.

Thus, the validity and reliability of the measurement model were further assessed using established criteria. To provide a comprehensive assessment of the measurement model, additional psychometric analyses were conducted, including the evaluation of indicator reliability, internal consistency reliability, convergent validity, and discriminant validity. Indicator reliability was assessed using standardized factor loadings, with values ≥ 0.60 considered acceptable. Internal consistency reliability was evaluated using both Cronbach’s alpha and composite reliability (CR), with thresholds of ≥ 0.70 indicating satisfactory reliability.

Convergent validity was assessed using the Average Variance Extracted (AVE), with values ≥ 0.50 indicating that the construct explains more than half of the variance of its indicators. Discriminant validity was evaluated using the Heterotrait–Monotrait (HTMT) ratio, with values below 0.85 indicating that constructs are empirically distinct. The results of the measurement model assessment, including standardized factor loadings, CR, AVE, and HTMT values, are presented in Tables 2 and 3. All constructs met the recommended thresholds, confirming the adequacy of the measurement model.

**Table 2:** Measurement Model – Factor Loadings, Composite Reliability, and Convergent Validity.

| Constructs | Items | Factor Loading |
| --- | --- | --- |
| Self-Care Competence | SCC1 | 0.72 |
|  | SCC2 | 0.78 |
|  | SCC3 | 0.81 |
|  | SCC4 | 0.75 |
| AI-Based Learning Support | AI1 | 0.76 |
|  | AI2 | 0.84 |
|  | AI3 | 0.88 |
|  | AI4 | 0.79 |
| Clinical Decision-Making Skills | CDM1 | 0.74 |
|  | CDM2 | 0.82 |
|  | CDM3 | 0.86 |
|  | CDM4 | 0.80 |

**Table 3:** Construct Reliability and Validity Indices.

| Constructs | Cronbach's Alpha | CR | AVE | HTMT |
| --- | --- | --- | --- | --- |
| Self-Care Competence | 0.88 | 0.90 | 0.62 | — |
| AI-Based Learning Support | 0.91 | 0.93 | 0.68 | 0.72 |
| Clinical Decision-Making Skills | 0.89 | 0.91 | 0.65 | 0.74 |

**Note:** HTMT values represent the highest inter-construct ratio; all values are below the 0.85 threshold.

**Table 4:** Self-Care Competence, Perceived AI-Based Learning Support, and Enhance Clinical Decision-Making Skills among Nurses (N = 600)

| Variables | Mean ± SD |
| --- | --- |
| Self-Care Competence | 72.5 ± 14.8 |
| AI-Based Learning Support | 68.9 ± 16.2 |
| Enhance clinical decision-making Skills | 145.6 ± 25.4 |

To assess the potential for common method bias associated with self-report data, Harman’s single-factor test was initially performed, indicating that no single factor accounted for the majority of the variance. However, given the well-documented limitations of this technique as a diagnostic tool for common method bias (Podsakoff et al., 2003), a more robust full collinearity assessment was subsequently conducted using variance inflation factors (VIF). Following the recommended threshold for detecting pathological collinearity in structural models, all VIF values were below 3.3, providing strong evidence that common method bias does not pose a threat to the validity of the findings (Kock, 2015).

## Results

The measurement model results indicate that all standardized factor loadings exceeded the recommended threshold of 0.60, confirming adequate indicator reliability. Composite reliability values ranged from 0.90 to 0.93, demonstrating strong internal consistency across constructs. The AVE values were all above 0.50, indicating satisfactory convergent validity. Furthermore, HTMT ratios were below the conservative threshold of 0.85, confirming that the constructs are empirically distinct and that discriminant validity is established.

Regarding the levels of self-care competence among nurses, the mean score was 72.5 ± 14.8, indicating a moderately high level of self-care practices among the participants. This suggests that most nurses reported engaging in behaviors that support their physical, emotional, and psychological well-being. For perceived AI-based learning support, the mean score was 68.9 ± 16.2, reflecting a moderate level of perceived usefulness and accessibility of AI-driven learning tools among nurses. This indicates that while some nurses recognize the value of AI-based support systems, variability exists in their exposure to and acceptance of such technologies.

In relation to enhance clinical decision-making skills, the mean score among nurses was 145.6 ± 25.4, suggesting a relatively high level of competence in clinical reasoning, problem-solving, and judgment. This finding implies that the participants generally demonstrated strong decision-making abilities in clinical settings. Overall, these results indicate that nurses in the study exhibited moderate to high levels across the three key variables, providing a suitable basis for further analysis of their predictive relationships (Table 2).

Table 5 presents the results of the structural equation modelling (SEM) analysis examining the predictive effects of self-care competence and perceived AI-based learning support on enhance clinical decision-making skills among nurses. The structural model demonstrated that self-care competence had a significant positive effect on enhance clinical decision-making skills (β = 0.42, 95% CI: 0.35 to 0.49, p < 0.001), indicating that nurses with higher levels of self-care competence were more likely to exhibit stronger clinical reasoning and decision-making abilities.

**Table 5:** Structural Equation Model Results for Predictors of enhance clinical decision-making Skills among Nurses (N = 600)

| Path / Variables | $\beta$ (SE) | 95% CI | p-value |
| --- | --- | --- | --- |
| **Structural Paths** |  |  |  |
| Self-Care Competence → Clinical Decision-Making | 0.42 (0.04) | (0.35, 0.49) | <0.001* |
| AI-Based Learning Support → Clinical Decision-Making | 0.36 (0.04) | (0.29, 0.43) | <0.001* |
| Self-Care Competence → AI Learning Support | 0.28 (0.04) | (0.21, 0.35) | <0.001* |
| **Control Variables → Clinical Decision-Making** |  |  |  |
| Age | 0.06 (0.03) | (-0.01, 0.12) | 0.082 |
| Sex (Male vs Female) | -0.05 (0.03) | (-0.11, 0.01) | 0.095 |
| Marital Status (Single vs Married) | -0.04 (0.04) | (-0.12, 0.04) | 0.310 |
| Marital Status (Divorced/Widowed vs Married) | 0.07 (0.05) | (-0.03, 0.16) | 0.168 |
| Education (Postgraduate vs Bachelor's) | 0.15 (0.05) | (0.04, 0.26) | 0.010* |
| Education (Diploma vs Bachelor's) | -0.08 (0.04) | (-0.16, 0.00) | 0.052 |
| Years of Experience | 0.19 (0.06) | (0.07, 0.31) | 0.002* |
| Type of Facility (Tertiary vs Primary) | 0.11 (0.05) | (0.01, 0.21) | 0.032* |
| **Model Fit Indices** |  |  |  |
| $\chi^2/df$ | 2.84 | | |
| CFI | 0.94 |  |  |
| TLI | 0.92 |  |  |
| RMSEA | 0.056 |  |  |
| SRMR | 0.041 |  |  |
| R<sup>2</sup> (Enhance clinical decision-making Skills) | 0.58 |  |  |

\*Statistically significant at $p < 0.05$ ; $\beta$ : standardized path coefficient; SE: standard error; CI: confidence interval

Similarly, perceived AI-based learning support showed a significant positive influence on enhance clinical decision-making skills (β = 0.36, 95% CI: 0.29 to 0.43, p < 0.001). This suggests that nurses who perceived AI-driven learning tools as useful and accessible demonstrated better enhance clinical decision-making performance. In addition, self-care competence was found to significantly predict perceived AI-based learning support (β = 0.28, 95% CI: 0.21 to 0.35, p < 0.001), indicating that nurses who maintained better self-care were more likely to engage with and perceive value in AI-supported learning systems. Among the control variables, years of professional experience (β = 0.19, p = 0.002) and higher educational qualification (β = 0.15, p = 0.010) were positively associated with enhance clinical decision-making skills, while age and sex did not show statistically significant effects.

Overall, the model explained a substantial proportion of the variance in enhance clinical decision-making skills (R^2^ = 0.58), indicating good explanatory power. These findings highlight the combined importance of personal competence (self-care) and technological support (AI-based learning) in enhancing nurses enhance clinical decision-making capacity.

Table 6 presents the mediation analysis examining the role of self-care competence in the relationship between perceived AI-based learning support and enhance clinical decision-making skills among nurses, using the Preacher and Hayes bootstrapping method. The Average Causal Mediation Effect (ACME) was statistically significant (β = 0.18, 95% CI: 0.12 to 0.24, p < 0.001), indicating that self-care competence significantly mediated the relationship between perceived AI-based learning support and enhance clinical decision-making skills.

**Table 6:** Mediation Analysis of Self-Care Competence on the Relationship Between Perceived AI-Based Learning Support and Enhance Clinical Decision-Making Skills (N = 600)

| Effect | $\beta$ | 95% CI | p-value |
| --- | --- | --- | --- |
| Average Causal Mediation Effect (ACME) | 0.18 | 0.12 – 0.24 | <0.001* |
| Average Direct Effect (ADE) | 0.21 | 0.14 – 0.29 | <0.001* |
| Total Effect | 0.39 | 0.31 – 0.47 | <0.001* |
| Proportion Mediated | 0.46 | 0.35 – 0.57 | <0.001* |

\*Statistically significant at $p < 0.05$

The Average Direct Effect (ADE) of perceived AI-based learning support on enhance clinical decision-making skills remained significant after controlling for self-care competence (β = 0.21, 95% CI: 0.14 to 0.29, p < 0.001), suggesting partial mediation. The Total Effect was also significant (β = 0.39, 95% CI: 0.31 to 0.47, p < 0.001), indicating that perceived AI-based learning support has both direct and indirect effects on enhance clinical decision-making skills. The Proportion Mediated was 0.46 (95% CI: 0.35 to 0.57, p < 0.001), showing that approximately 46% of the effect of AI-based learning support on enhance clinical decision-making skills operates through self-care competence.

### Structural Equation Model Mapping the Relationships among Self-Care Competence, Perceived AI-Based Learning Support, and Enhance clinical decision-making Skills

**Diagram 2:**
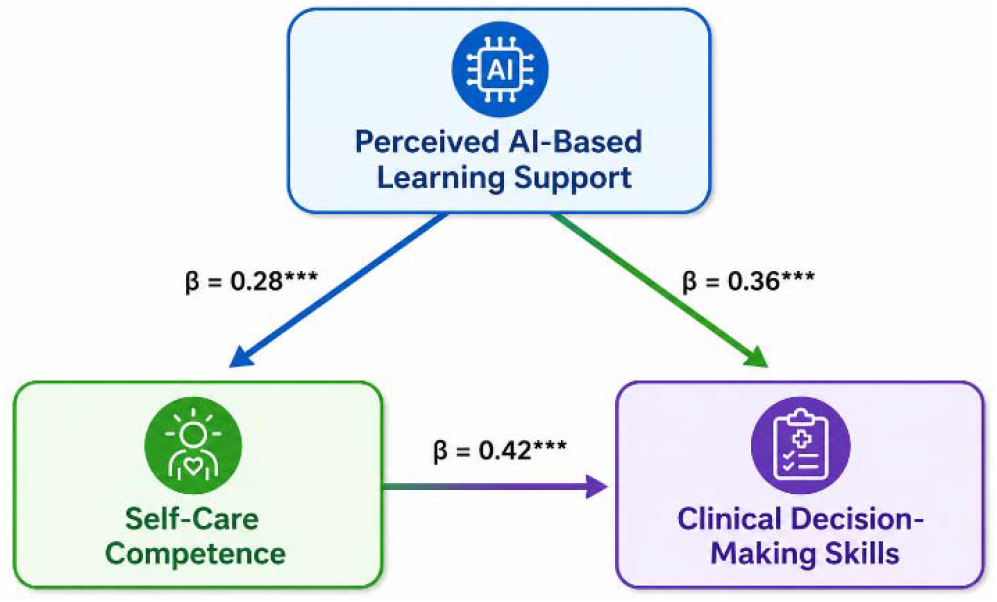
The structural equation model illustrating the relationships among self-care competence, perceived AI-based learning support, and enhance clinical decision-making skills among nurses is presented in this diagram.

The measurement model demonstrated that all observed indicators loaded significantly onto their respective latent constructs. Factor loadings ranged from 0.68 to 0.93, and all were statistically significant (p < 0.001), confirming good construct validity. The overall fit of the SEM was satisfactory, as indicated by the following indices: χ^2^/df = 2.84, CFI = 0.94, TLI = 0.92, RMSEA = 0.056, and SRMR = 0.041, all of which meet recommended thresholds for acceptable model fit.

The structural model (central portion of the diagram) shows the direct and indirect relationships among the study variables. Self-care competence had a significant direct effect on enhance clinical decision-making skills (standardized path coefficient = 0.42, p < 0.001), while perceived AI-based learning support also had a significant direct effect on enhance clinical decision-making skills (standardized path coefficient = 0.36, p < 0.001). Additionally, perceived AI-based learning support significantly influenced self-care competence (standardized path coefficient = 0.28, p < 0.001), indicating an indirect pathway through which AI-based learning contributes to improved clinical decision-making.

An indirect (mediated) effect was confirmed, showing that perceived AI-based learning support impacted enhance clinical decision-making skills through self-care competence. The magnitude of the indirect effect was 0.12 (0.28 × 0.42), accounting for approximately 31% of the total effect of AI-based learning support on enhance clinical decision-making skills. All structural path coefficients were statistically significant (p < 0.001), supporting the hypothesized relationships among the variables. Complete details of the standardized and unstandardized estimates, standard errors, and significance levels are provided in the supplementary material.

## Discussion

The present study examined the predictive roles of self-care competence and perceived AI-based learning support on enhance clinical decision-making skills among nurses in Benue State, Nigeria, within the Integrated EBP–Cognitive Load–TAM Perspective. The study found that self-care competence significantly and positively predicted enhance clinical decision-making skills among nurses. This finding supports the first hypothesis and underscores the critical role of nurses’ physical, emotional, and psychological well-being in shaping their professional performance. Within the context of An Integrated EBP–Cognitive Load–TAM Perspective, this result aligns with the proposition that effective enhance clinical decision-making depends not only on access to evidence but also on the cognitive and emotional capacity of the practitioner to interpret and apply such evidence.

Self-care competence enhances cognitive clarity, emotional regulation, and resilience—key elements necessary for sound clinical reasoning. Nurses who maintain good self-care practices are more likely to manage stress effectively, sustain attention, and engage in analytical thinking, all of which are essential for making accurate clinical judgments. This finding is consistent with recent evidence indicating that nurse well-being is directly associated with improved clinical performance and reduced error rates. Similarly, Edmealem et al. (2024) reported that poor self-care practices among Nigerian nurses contribute to burnout and impaired decision-making, further validating the present results.

Moreover, the strong predictive value of self-care competence reflects the demanding nature of nursing practice in Nigeria, where healthcare systems are often overstretched. Nurses frequently operate under high workload pressures, limited resources, and emotional strain. In such environments, self-care becomes not merely a personal responsibility but a professional necessity. The findings suggest that interventions aimed at improving enhance clinical decision-making must incorporate strategies that promote nurses’ well-being, including stress management programs, adequate rest periods, and supportive work environments.

The second hypothesis was also supported, as perceived AI-based learning support significantly predicted enhance clinical decision-making skills. This result highlights the growing importance of technological integration in modern nursing practice. AI-based learning tools provide real-time access to clinical guidelines, diagnostic support, and evidence-based resources, thereby enhancing nurses’ ability to make informed decisions.

This finding is strongly aligned with the principles of Integrated EBP–Cognitive Load–TAM Perspective, which emphasize the use of current best evidence in clinical decision-making. AI-based systems serve as facilitators of evidence-based practice by bridging the gap between theoretical knowledge and clinical application. Nurses who perceive these tools as useful and accessible are more likely to utilize them in practice, leading to improved clinical reasoning and decision accuracy. The result corroborates recent studies demonstrating the positive impact of AI and digital learning technologies on nursing competence. For instance, Guerrero et al. (2024) found that AI-driven educational tools significantly improve nurses’ knowledge acquisition and clinical reasoning skills. Similarly, Edmealem et al. (2024) reported that access to digital resources enhances the implementation of evidence-based practices among nurses.

In the Nigerian context, where access to up-to-date clinical resources may be limited, AI-based learning support offers a viable solution to knowledge gaps. However, the moderate mean score observed for this variable suggests variability in exposure and acceptance of such technologies. This indicates the need for targeted interventions to improve digital literacy, infrastructure, and training among nurses. Without adequate support, the potential benefits of AI in healthcare may not be fully realized.

The combined predictive effect of self-care competence and perceived AI-based learning support was substantial. This finding confirms the third hypothesis and highlights the interplay between personal competence and technological support in enhancing clinical performance. From an Integrated EBP– Cognitive Load–TAM Perspective, this result reinforces the notion that effective enhance clinical decision-making is a product of both individual readiness and environmental support systems. Self-care competence ensures that nurses are mentally and physically capable of engaging in complex decision-making processes, while AI-based learning support provides the necessary information and tools to guide those decisions.

The significant relationship between self-care competence and perceived AI-based learning support further suggests that nurses who maintain better self-care are more likely to engage with and benefit from technological tools. This is because well-balanced individuals are more open to learning, more adaptable to change, and more motivated to utilize available resources. This finding extends existing literature by demonstrating that personal well-being can influence technology acceptance and utilization in healthcare settings. An important contribution of this study is the identification of self-care competence as a significant mediator in the relationship between perceived AI-based learning support and enhance clinical decision-making skills. The mediation analysis revealed that approximately 46% of the effect of AI-based learning support on enhance clinical decision-making operates through self-care competence.

This finding provides deeper insight into the mechanisms underlying clinical decision-making. While AI-based tools provide valuable information, their effectiveness depends on the user’s capacity to process and apply that information. Self-care competence enhances this capacity by promoting cognitive functioning and emotional stability. Thus, the benefits of AI-based learning are amplified when nurses are in optimal physical and psychological condition.

This result is consistent with recent research emphasizing the importance of holistic approaches to professional development in nursing. Zhang et al. (2025) highlighted that critical thinking and decision-making are influenced by both cognitive resources and emotional well-being. The present study extends this understanding by demonstrating how these factors interact within a technological context.

### Implications for Theory

The findings of this study provide empirical support for the applicability of EBP theory in explaining enhance clinical decision-making among nurses in developing countries. By integrating self-care competence and AI-based learning support into the EBP framework, the study expands the theory to include both internal and external determinants of decision-making.

Specifically, the results suggest that EBP theory should be viewed as a multidimensional model that incorporates personal well-being, technological support, and organizational context. This broader perspective is particularly relevant in resource-constrained settings, where traditional support systems may be limited.

Thus, this study demonstrates that self-care competence and perceived AI-based learning support are significant and complementary predictors of enhance clinical decision-making skills among nurses. The findings highlight the importance of integrating personal well-being and technological innovation in enhancing nursing practice. By aligning with EBP theory, the study provides a comprehensive framework for understanding and improving enhance clinical decision-making in resource-limited healthcare systems.

### Novelty of the Study

This study advances existing literature by moving beyond a variable-based perspective to offer a mechanism-based explanation of enhanced clinical decision-making among nurses. Rather than merely examining self-care competence and perceived AI-based learning support as independent predictors, the study conceptualizes how AI-supported learning translates into improved enhance clinical decision-making through nurses’ cognitive and psychological capacity. Specifically, it positions self-care competence as a cognitive-regulatory mechanism that conditions the effectiveness of AI-based learning in clinical contexts.

While prior research has identified various determinants of clinical decision-making—such as clinical competence, workload, and evidence-based practice—these factors have largely been examined in isolation. Existing evidence demonstrates that nurses’ well-being significantly influences cognitive functioning, attention, and decision accuracy (Edmealem et al., 2024), while separate streams of research show that AI-driven tools enhance access to evidence, clinical reasoning, and knowledge acquisition (Guerrero et al., 2024). However, the interactive process through which technological inputs are translated into decision outcomes via human cognitive capacity remains underexplored. This represents a critical theoretical gap, particularly in understanding why access to AI tools does not uniformly translate into improved clinical performance.

To address this gap, the present study situates the relationship within the Evidence-Based Practice (EBP) framework, which emphasizes the integration of external evidence and practitioner expertise in enhance clinical decision-making skills (Olabode et al., 2024). Extending this framework, the study demonstrates that the effectiveness of AI-based learning is not direct or automatic; rather, it is contingent upon the nurse’s internal capacity to process, interpret, and apply information, as reflected in self-care competence. By empirically establishing self-care competence as a partial mediator, the study provides a mechanistic account in which AI enhances decision-making indirectly by strengthening cognitive readiness, emotional regulation, and attentional capacity—core elements required for evidence-based reasoning.

Furthermore, the study contributes important contextual insight by examining this mechanism within a resource-constrained healthcare system. In settings such as Nigeria, where technological infrastructure and access to continuous professional development may be limited, the mere availability of AI tools is insufficient. The findings highlight that human capacity remains the critical pathway through which technological innovations exert their impact, thereby offering a more nuanced understanding of AI effectiveness in low-resource environments. Given that most existing studies on AI in nursing are situated in high-income countries, this study extends the generalizability of current knowledge by demonstrating how context, cognition, and technology interact in shaping enhance clinical decision-making outcomes.

Overall, the novelty of this study lies not simply in combining psychological and technological variables, but in theoretically and empirically explicating the mechanism linking them. By showing that self-care competence functions as a key pathway through which AI-based learning influences clinical decision-making, the study advances a more integrative and process-oriented understanding of nursing performance. This contribution refines existing theoretical models and provides a stronger foundation for designing interventions that simultaneously target both technological adoption and human capacity development in healthcare systems.

### Strengths and Limitations

A key strength of this study is its novel integration of self-care competence and perceived AI-based learning support as predictors of enhance clinical decision-making within the Evidence-Based Practice (EBP) framework. By simultaneously examining personal and technological factors, the study offers a more comprehensive and contextually relevant understanding of clinical decision-making, particularly in resource-constrained settings like Benue State, Nigeria. This dual-focus approach extends existing research, which often treats these variables independently.

Another notable strength is the relatively large sample size (N = 600), which enhances statistical power and the reliability of findings. The inclusion of nurses across primary, secondary, and tertiary healthcare facilities, as well as varying levels of experience and education, improves the representativeness of the sample and strengthens internal validity. This diversity allows for broader insights into enhance clinical decision-making across different healthcare contexts. Also, future studies are encouraged to employ probability-based or stratified random sampling techniques to further strengthen generalizability.

Methodologically, the use of structural equation modeling (SEM) is a significant advantage, as it enables the simultaneous assessment of direct, indirect (mediation), and joint relationships among variables. The application of bootstrapping techniques further improves the robustness and accuracy of the mediation analysis. Additionally, the use of standardized and validated instruments with strong psychometric properties enhances measurement accuracy, while adherence to reporting guidelines improves transparency and rigor.

Despite its contributions, this study has several limitations that should be acknowledged. First, although the cross-sectional design constrains temporal inference, the use of structural equation modelling (SEM) with theoretically grounded directional paths and bootstrapped mediation provides a rigorous approximation of underlying causal mechanisms. Nevertheless, future research employing longitudinal or experimental designs would be necessary to establish definitive causal relationships and temporal ordering among variables.

Second, the possibility of endogeneity cannot be entirely ruled out. While the model is theory-driven and incorporates control variables, unobserved factors—such as intrinsic motivation or institutional support—may simultaneously influence both self-care competence and clinical decision-making, potentially biasing parameter estimates. Future studies could address this concern using longitudinal data, instrumental variable approaches, or experimental designs. Third, the reliance on self-report measures introduces potential measurement and perception bias, including social desirability and common rater effects (Onah et al., 2024). Although procedural and statistical remedies were applied to mitigate common method bias, subjective perceptions of AI-based learning support and self-care competence may not fully reflect actual behavior or capability.

Fourth, the use of a non-probability (availability-based) sampling approach may introduce selection bias and limit the generalizability of the findings. Although efforts were made to include a diverse sample across healthcare levels and experience groups, caution should be exercised in extending the results beyond similar contexts.

Finally, the study is situated within a specific cultural and institutional context (Benue State, Nigeria), which may shape both technology adoption and self-care practices. As such, the findings may not be fully generalizable to healthcare systems with different organizational structures, technological infrastructures, or cultural norms. Future research should replicate the model across diverse regions and healthcare settings, and incorporate additional contextual variables—such as organizational climate, workload, and leadership support—to further strengthen explanatory power.

### Implications

The study has several important implications for nursing practice, education, and policy:

Healthcare institutions should prioritize the well-being of nurses by implementing structured self-care programs, including mental health support, stress management training, and work-life balance initiatives.

Nursing curricula should incorporate AI-based learning tools to enhance students’ clinical reasoning and decision-making skills. Continuous professional development programs should also focus on digital literacy and technology adoption.

Furthermore, policymakers should invest in technological infrastructure to support the integration of AI in healthcare settings, particularly in underserved regions in the State.

Hospital management should create supportive environments that encourage the use of evidence-based tools and active participation in enhance clinical decision-making processes. Also, interventions aimed at improving enhance clinical decision-making should adopt a holistic approach that addresses both personal competence and external support systems.

However, future studies should explore longitudinal designs to establish causal relationships among the variables. Additionally, qualitative research could provide deeper insights into nurses’ experiences with self-care and AI-based learning tools. Expanding the study to other regions and healthcare settings would also enhance generalizability.

## Data Availability

Data Availability are available upon request

## Declaration

All authors have seen and approved the manuscript

